# Convergent multidimensional neural phenotypes in human patients and schizophrenia-relevant mouse models identify reduced recurrent excitation in cortical circuits

**DOI:** 10.64898/2026.09.22.26363726

**Authors:** Kenneth Wengler, Jordan P. Hamm, Yuriy Shymkiv, Kenneth D. Miller, Rafael Yuste, Guillermo Horga

**Author notes:** These authors contributed equally (Rafael Yuste, Guillermo Horga). **Correspondence:** Kenneth Wengler, PhD, Department of Psychiatry, Icahn School of Medicine at Mount Sinai 100 E 104^th^ St, 3^rd^ Floor Room 305F, New York, NY 10029. **Previous Presentation:** This work was presented in a symposium at the Society of Biological Psychiatry Annual Meeting in New York, NY on 4/30/2026.

## Abstract

**Objective:** NMDA-receptor hypofunction has been implicated as a core pathophysiological mechanism in schizophrenia, but conflicting evidence exists regarding the locus of the hypofunction—i.e. predominantly affecting receptors on pyramidal neurons or interneurons. To investigate the locus of NMDA-receptor hypofunction we integrated imaging data from humans and mice with biophysical model simulations.

**Methods:** Translational imaging measures—intrinsic neural timescales (INT) and spontaneous co-activation pattern similarity—were investigated in three schizophrenia-relevant mouse models (chronic ketamine administration, n=6; *Df(16)A*^+/−^, n=7; and *SETD1A*^+/-^, n=9) and matched control mice (n=5; n=7; and n=8, respectively) using two-photon calcium imaging, and individuals with schizophrenia (n=127) and matched non-clinical controls (n=152) using functional MRI. The stabilized supralinear network biophysical model was used to simulate NMDA-receptor hypofunction and its impact on the translational imaging measures.

**Results:** We identified convergent neural phenotypes of shorter intrinsic neural timescales (INT) and reduced spontaneous co-activation pattern similarity in both the schizophrenia-relevant mouse models and the individuals with schizophrenia compared to matched controls. *In silico* simulations using the stabilized supralinear network biophysical model recapitulated the convergent neural phenotypes through reduced excitatory-to-excitatory connection strength (i.e. NMDA-receptor hypofunction on pyramidal neurons) and predicted that INT and pattern similarity are correlated; this prediction was confirmed in both the mouse and human data.

**Conclusions:** These results identify NMDA-receptor hypofunction on pyramidal neurons as a general and translationally valid mechanism for NMDA-receptor hypofunction in schizophrenia. These convergent translational findings suggest prioritizing the development of treatments for schizophrenia that increase recurrent excitation.

## INTRODUCTION

Schizophrenia is a serious mental illness present in ∼1% of the global population (1). Despite the development of antipsychotics in the 1960’s for treating the positive symptoms of schizophrenia (i.e. hallucinations and delusions) (2, 3), treatments for other symptom domains (i.e. negative symptoms and cognitive deficits) have been limited (4, 5). While substantial efforts have been made, a translational gap remains in the treatment-development process due to unclear pathophysiology of the disease (6, 7).

A critical challenge in studying the pathophysiology of schizophrenia is the notably diverse etiology and high degree of heterogeneity with respect to symptoms and genetic causes (8). Genetic data implicate alterations in excitatory synapses with both common and rare variants converging on N-methyl-D-aspartate (NMDA)-receptor function (e.g., GRIN2A) (9, 10) and the largest number of differentially expressed genes present in excitatory neurons (11). NMDA-receptor hypofunction as a core component of schizophrenia is further supported by pharmacological findings that NMDA-receptor antagonists (e.g., ketamine and phencyclidine) cause symptoms indistinguishable from idiopathic schizophrenia (12, 13) and exacerbate symptoms in individuals with schizophrenia (14, 15). Furthermore, post-mortem studies have consistently identified reduced dendritic spines (the site of excitatory inputs) on pyramidal cells (16–18), and a recent meta-analysis reported brain-wide reductions in the number of excitatory synapses in schizophrenia (19). Despite substantial evidence supporting NMDA-receptor-related pathology in schizophrenia, the locus of the hypofunction (i.e. predominantly affecting receptors on pyramidal neurons or interneurons) is widely debated. While the functional consequence—e.g., working memory impairment—of reduced excitation (NMDA-receptor hypofunction on pyramidal neurons) or disinhibition (NMDA-receptor hypofunction on interneurons) may be similar (20–22), the determination of the locus of NMDA-receptor hypofunction could have profound implications for treatment targets.

Biophysical models—recurrent neural networks of excitatory and inhibitory neurons with biologically realistic parameters—allow for theoretical investigation of NMDA-receptor hypofunction (23, 24). Some recent biophysical modelling work suggests NMDA-receptor hypofunction in schizophrenia predominantly affects interneurons due to the model’s ability to recapitulate some (20, 25)—but not all (26–29)—aspects of behavioral abnormalities during working memory tasks when reducing excitatory-to-inhibitory neuron connection strengths. Conversely, previous biophysical modelling work has recapitulated several electroencephalogram (EEG) and functional magnetic resonance imaging (fMRI) findings via NMDA-receptor hypofunction on pyramidal neurons (i.e. when reducing excitatory-to-excitatory neuron connection strengths) (30, 31).

To arbitrate between alternative hypotheses of the locus of NMDA-receptor hypofunction in schizophrenia, it is paramount to utilize convergent findings across preclinical and clinical studies, as well as biophysical model findings that are convergent across multiple phenotypes. Here we: 1) leverage the replicated finding of reduced neural timescales (INT) in individuals with schizophrenia (30, 32–34) to test for backward-translational convergence in three schizophrenia-relevant mouse models; 2) leverage the convergent findings from the three schizophrenia-relevant mouse models showing reduced similarity of spontaneous neuronal co-activation patterns (35, 36) to test for forward-translational convergence in individuals with schizophrenia; 3) propose a biophysical-modeling framework that recapitulates the convergent multidimensional phenotypes through NMDA-receptor hypofunction; and 4) test predictions of the biophysical model in the mouse and human data.

## METHODS

### Schizophrenia Combined Dataset

All human data presented here come from datasets published in Wengler et al. (2020) (30). As previously described (30), T1w images and rs-fMRI data were obtained for 331 non-clinical control subjects and 254 individuals diagnosed with schizophrenia from four publicly available datasets. Three of these datasets were from the SchizConnect repository (BrainGluSchi (37), COBRE (38, 39), and NmorphCH (40)) and one was from the OpenfMRI repository (UCLA (41)). The final sample after quality-control checks (see Supplement for details) consisted of 127 individuals with schizophrenia and 152 age- and gender-matched controls (Supplementary Table S1).

Resting-state fMRI (rs-fMRI) data were collected for each subject during an eyes-open-on-fixation session and preprocessed using a standardized AFNI pipeline (see Supplement for details). The preprocessed rs-fMRI data were then postprocessed according to recent recommendations(42) (see Supplement for details).

INT maps were calculated from the postprocessed rs-fMRI data as previously described (30) (see Figure 1 for overview). Briefly, the autocorrelation function of the rs-fMRI signal at each voxel was estimated and the sum of the autocorrelation coefficients during the initial positive period was calculated and multiplied by the repetition time. Voxelwise INT maps were parcellated into 180 cortical parcels using HCP-MMP1.0 in volumetric space averaging across the left and right hemispheres. See Supplement for further details on INT estimation.

**Figure 1.**
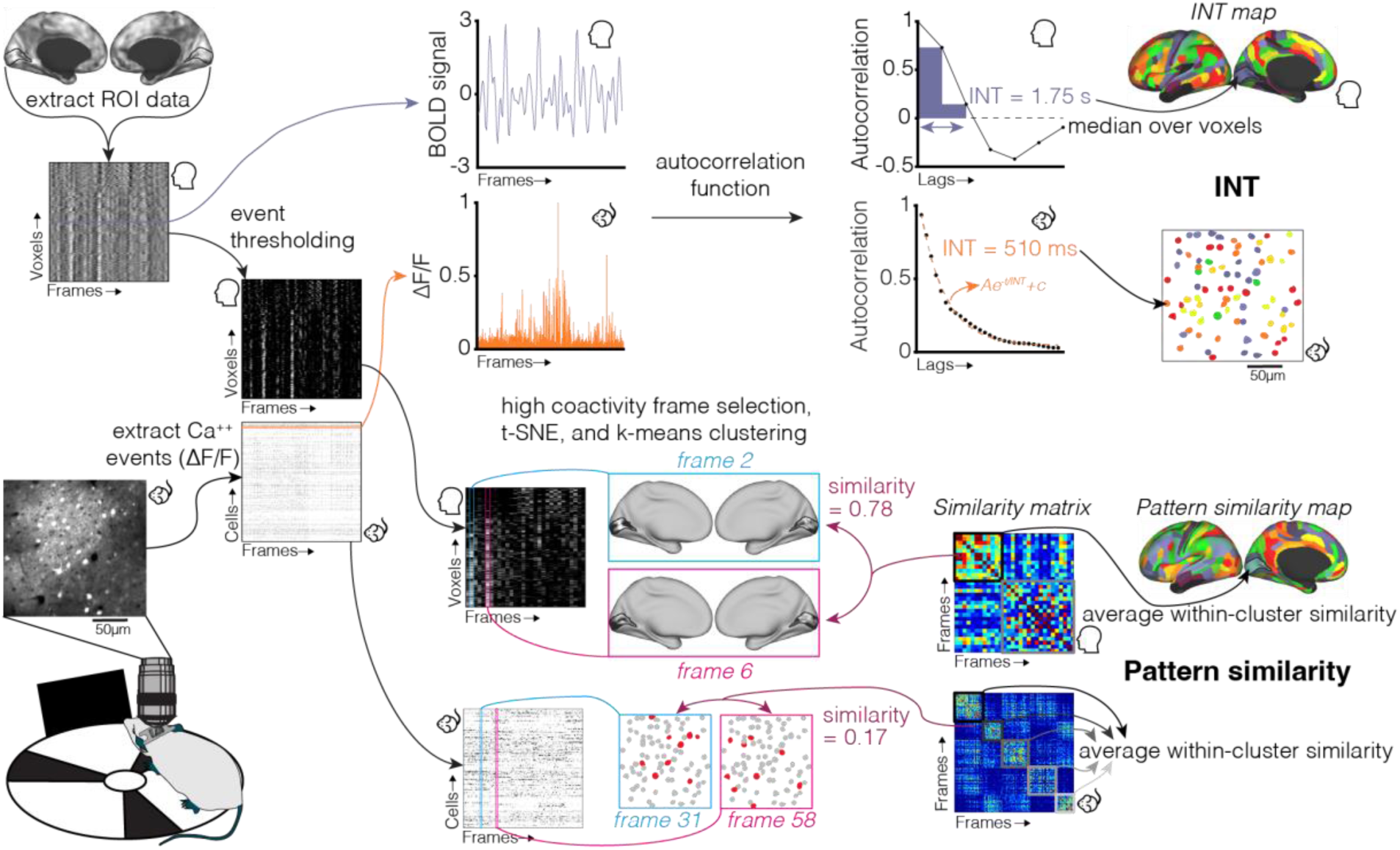
Overview of rs-fMRI and calcium imaging analysis pipeline. Rs-fMRI data were collected in humans during the eyes open condition. 2p-Ca^++^ imaging data were collected in mice viewing a blank monitor while on a treadmill; imaging was performed in layer-2/3 of V1. **For the human rs-fMRI data.** For each region of interest (ROI; V1 shown here) the rs-MRI timecourses of all voxels within the ROI are extracted and temporally z-scored after standard preprocessing. For intrinsic neural timescale (INT) estimation, the autocorrelation function of each voxel’s timecourse is calculated and the area under the curve of the initial positive period is taken as the voxel’s INT. This is repeated for all voxels within the ROI and the median timescale over all voxels is taken as the ROI’s INT. This is then repeated for all ROIs. For pattern similarity estimation, neural events are first defined by thresholding the BOLD signal (values <=1 are set to 0); the 30 frames with the highest level of coactivity over all voxels are then selected and t-distributed stochastic neighbor embedding (t-SNE) followed by k-means clustering (with two clusters) is carried out; the cosine similarity is subsequently calculated for all frame-frame pairs (*Similarity matrix*) and the average similarity within the dominant cluster (i.e. with greater within-cluster similarity) is taken as the ROI’s pattern similarity. This is then repeated for all ROIs. **For the mouse 2p-Ca^++^ imaging data.** Standard image processing is performed to identify individual cells from layer-2/3 in V1 and Ca^++^ events are extracted using the first discrete derivative normalized by the within-cell maximum to generate Δf/f timecourses. For INT estimation, the autocorrelation of each cell’s timecourse is calculated and fit with an exponential decay curve and the decay rate is taken as the cell’s INT. This is repeated for all cells and the median INT over all cells is taken as the mouse’s INT. For pattern similarity estimation, high coactivity frames that exceeded the 99.9^th^ percentile of coactivity from time-shuffled datasets are selected and t-SNE followed by k-means clustering (with number of clusters equal to number of cells divided by 33) is carried out; the Pearson correlation is subsequently calculated for all frame-frame pairs (*Similarity matrix*) and the average similarity within the clusters normalized by the average similarity of all frame pairs is taken as the mouse’s pattern similarity.

Pattern similarity maps were calculated from the postprocessed rs-fMRI data according to methods similar to those used for 2p-Ca^2+^ imaging data with neural events being defined as in co-activation pattern analyses of rs-fMRI data (43, 44). Pattern similarity values for each of the 180 cortical parcels were estimated (see Figure 1 for overview) through event thresholding, high-coactivity frame selection, k-means clustering of dimensionality reduced data and within-cluster cosine similarity (c_WC_). The c_WC_ value within the dominant cluster (i.e. the cluster with higher c_WC_) was taken as the pattern similarity. See Supplement for further details on pattern similarity estimation.

The parcellated INT and pattern similarity maps were then harmonized using ComBat (45–49) controlling for age and sex. The harmonization parameters were estimated using data from the non-clinical controls and subsequently applied to all subjects to minimize the potential removal of biological variability between non-clinical controls and individuals with schizophrenia (50–52).

### Mouse Models of Schizophrenia-Relevant Disease Mechanisms Combined Dataset

All animal work was carried out under IACUC approved protocols at Columbia University. All mouse data presented here come from image-processed datasets published in Hamm et al. (2017) (35) and Hamm et al. (2020) (36). All analyses are focused on brain activity collected in a darked room in awake mice in the absence of structured or changing visual input, which we refer to herein as “spontaneous activity.” Pattern similarity analyses match those described in Hamm et al. (2020) (36), and data from Hamm et al. (2017) (35) were re-analyzed to match those procedures. INT was not previously analyzed in either of these datasets. The surgical, experimental, and data processing steps are identical to those works (35, 36) and are only detailed briefly here.

Three previously published mouse datasets were used: a group of mice given 100mg/kg/day ketamine (n=6; 4 males) or saline (n=5; 3 males) continuously for a week via a subcutaneous minipump (from Hamm et al., 2017 (35)); a group of *Df(16)A*^+/-^ mice (mirroring the 22q11.2 microdeletion syndrome; n=7, all male) or their littermate/cagemate wild-type (WT) controls (n=7, all male; from Hamm et al., 2017 (35)); or a group of *SETD1A*^+/-^ mice (n=9; all male) or their littermate/cagemate WT controls (n=8; all male; from Hamm et al., 2020 (36)). All mice were aged between P84 and P180 during recordings.

Two-photon calcium imaging was performed in layer 2/3 of left primary visual cortex (V1). carried out in a dark room in the absence of visual stimulation. The activity of cortical neurons was recorded by imaging fluorescence (F) changes under a two-photon microscope (Bruker, Billerica, MA). For the *SETD1A*^+/-^ recordings, images were collected at ≈10 frames per second; for the ketamine and *Df(16)A*^+/-^ recordings, images were collected at ≈3.4 frames per second. Imaging consisted of a visual stimulation condition (15 minutes), followed by 20–40 minutes of awake rest in a dark room with the monitor off, followed by a second visual stimulation. The raw images were processed using standardized procedures (see Supplement for details). The processed F traces were then filtered with a 1-second LOWESS envelope and the first discrete derivative was scored as ΔF (for the INT analysis) or within-cell maximum normalized ΔF for population analyses (for the pattern similarity analysis).

Individual neuron intrinsic timescales were calculated by fitting an exponential decay function to the ΔF timeseries (53) and the mouse’s INT was taken as the median of the decay rates across the imaged neurons (see Figure 1 for overview). See Supplement for further details on INT estimation.

Pattern similarity of spontaneous ensemble activations during resting state was estimated as reported in Hamm et al. (2020) (36) (see Figure 1 for overview) and included bootstrapping to identify true “ensemble activations,” k-means clustering of dimensionality reduced ensemble activations, and within-cluster Pearson correlation similarity normalized by the average unclustered correlation value across all ensemble activations (r_WCN_). The average r_WCN_ value across clusters was taken as the mouse’s pattern similarity. See Supplement for further details on pattern similarity estimation.

### Statistical Analysis

Linear models were used to test for group differences in INT and pattern similarity, and for relationships between INT and pattern similarity across subjects/mice. Human subject analyses controlled for age, sex, head motion (mean framewise displacement, percent of censored frames, and the square of percent of censored frames (42)). Mouse analyses controlled for dataset (i.e. chronic ketamine, *Df(16)A*^+/-^, and *SETD1A*^+/-^) and control analyses included interactions with dataset. Analyses were considered significant at an alpha of 0.05 (two-tailed) and were conducted using MATLAB, version R2024a.

### Biophysical Model

We utilized the stabilized supralinear network (SSN) (54–58) with different synaptic receptor types(56). The SSN is highly constrained by biological properties and has been shown to recapitulate many features of the V1. Here, we implemented a retinotopically-structured SSN model of V1 that models the cortex as a two-dimensional grid with an *E* and *I* sub-population (corresponding to SSN units) at each grid location, corresponding to a cortical column. The grid two-dimensional grid covered an area of 6.4 mm × 6.4 mm and contained 17 × 17 evenly spaced cortical columns (each containing and *E* and *I* sub-population) such that the total number of *E* and *I* populations is 289. We account for the distinct dynamics of currents through different synaptic receptor channels: AMPA, GABA_A_ (henceforth GABA), and NMDA.

To simulate the resting-state condition (i.e. spontaneous activity), we input spatially correlated and identically distributed noise across our excitatory units, and took it to be temporally correlated pink noise with zero mean and unit variance. The degree of noise correlation between neurons was decayed as an exponential function of distance between neurons with a decay constant of 0.4 mm (one grid interval) (59–61). Biophysical model simulations were run using the forward Euler method with a time step of 1 ms and a total simulation time of 10 minutes. Simulations were performed while independently varying one of each of the neuronal connectivity strengths (*W_EE_, W_IE_, W_EI_,* or *W_II_*) while keep all other parameters fixed. The connectivity strengths were varied by factors ranging from 0.9 to 1.1 (i.e. 10% decrease to 10% increase) in steps of 5% (i.e. 5 separate values), for a total of 20 separate model simulations (i.e. 5 connectivity values for each of the 4 connectivity parameters). See Supplement for further details on biophysical model simulations.

The simulated firing rates from each of the 289 excitatory neurons were used to calculate INT and pattern similarity (to match the 2P-Ca^2+^ data that imaged layer 2/3 pyramidal neurons). The first minute of simulated firing rates were discarded and the simulated firing rates were downsampled to a temporal resolution of 10 Hz. The model’s INT was estimated using the same procedure as the calcium imaging data. The model’s pattern similarity was estimated using the same procedure as the calcium imaging data using the top 200 timepoints (determined via bootstrapping of the unperturbed model) and k=5 (determined via maximizing the silhouette index of the ensemble activations from the unperturbed model).

## RESULTS

### Convergent deficits in both INT and pattern similarity

Given several reports of shorter INT in individuals with schizophrenia (30, 32–34), we reanalyzed resting-state 2P-Ca^2+^ data from three mouse models of schizophrenia-relevant disease mechanisms [*Df*(*16*)*A*^+/-^ (n=7; n=7 controls); *SETD1A*^+/-^ (n=9; n=8 controls); and chronic ketamine (n=6; n=5 controls)] to calculate INT. We found that V1 INT was significantly shorter in the mouse models compared to the control mice (t=-2.10, p=0.043; Figure 2a); results were consistent across mouse models (all interaction effects p>0.511). We also reanalyzed rs-fMRI data from individuals with schizophrenia (n=127) and non-clinical controls (n=152) and found (as expected based on previous analysis of these data(30) but now using optimized methods for handling potential confounds from respiration and head motion (42)) shorter whole-brain averaged INT (t=-2.34, p=0.020; Figure 2b). Next, we estimated the pattern similarity of spontaneous activations using the rs-fMRI data from the individuals with schizophrenia and non-clinical controls and found reduced whole-brain averaged pattern similarity in the individuals with schizophrenia (t=-2.62, p=0.009; Figure 2d). Similarly, when combining the mouse 2P-Ca^2+^ data, we found (as expected based on previous analyses of these data (35, 36)) reduced pattern similarity of V1 spontaneous activity in the mouse models (t=-4.04, p<0.001; Figure 2c); results were consistent across mouse models (all interaction effects p>0.487). See Supplement (Supplementary Figure S1) for group-average cortical maps of INT and pattern similarity for individuals with schizophrenia and non-clinical controls, a cortical map of the group comparisons (i.e. parcel-wise analysis), and results for human V1 which showed similar effects.

**Figure 2.**
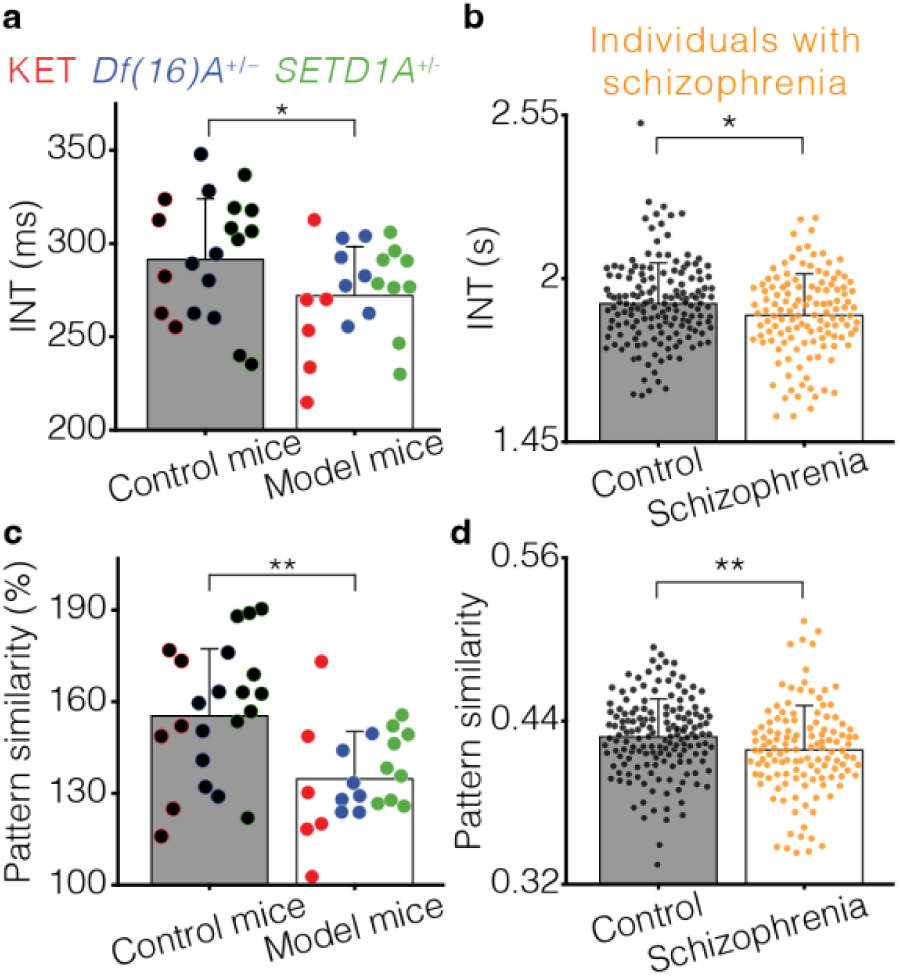
INT and pattern similarity deficits across mouse models of schizophrenia-relevant disease mechanisms and individuals with schizophrenia. **a.** INT is significantly shorter in V1 of mouse models of schizophrenia-relevant disease mechanisms compared to wild-type controls. **b.** As previously reported, whole-brain INT is significantly shorter in individuals with schizophrenia compared to non-clinical controls. **c.** As previously reported, pattern similarity is significantly lower in V1 of mouse models of schizophrenia-relevant disease mechanisms compared to wild-type controls. **d.** Whole-brain pattern similarity is significantly lower in individuals with schizophrenia compared to non-clinical controls. *, p<0.05; **, p<0.01.

### Biophysical model recapitulates INT and pattern similarity deficits

To explore the circuit-level mechanism of pattern similarity and INT deficits, we utilized the stabilized supralinear network (SSN), a highly-constrained, state-of-the-art, and well-validated biophysical model— a network of excitatory I and inhibitory (I) neurons with biologically realistic properties—of V1 (54–58). A retinotopically-structured SSN model of V1 (Figure 3a) was simulated with noise input to each neuron to model spontaneous (i.e. resting state) activity. The SSN produced modular patterns of activity consistent with observed ensemble activity (Figure 3b). As previously demonstrated (54–58), the SSN accounts for a broad range of canonical cortical circuit properties including normalization (62, 63) (Figure 3c–e), surround suppression (64, 65) (Figure 3f), and contrast dependence of gamma oscillations (66) (Figure 3g). To arbitrate between alternative hypotheses of the locus of NMDA-receptor hypofunction in schizophrenia, the SSN was simulated while independently varying the inter-neuronal connection strengths from E→E cells (Figure 3h) and E→I cells (Figure 3i). Notably, reductions in E→E connection strengths were able to recapitulate the reductions of INT and pattern similarity observed in individuals with schizophrenia and the mouse models, while reductions in E→I connection strengths lead to increased INT and pattern similarity. This effect likely arises due to stronger E→E connections causing greater self-amplification of activity patterns resulting in stronger (i.e. higher pattern similarity) and longer lasting (i.e. longer INT) patterns. Meanwhile, increased E→I likely suppresses these patterns leading to weaker and more quickly decaying patterns. See Supplement (Supplementary Figure S2) for model simulations varying other inter-neuronal connections and simulations investigating stimulus-evoked gamma power in which reductions—also recapitulated with reduced E→E connection strength—were previously reported across all three mouse models (35, 36) and consistently observed in individuals with schizophrenia (67) although it was not assessed in the current sample of individuals with schizophrenia (requisite data not collected).

**Figure 3.**
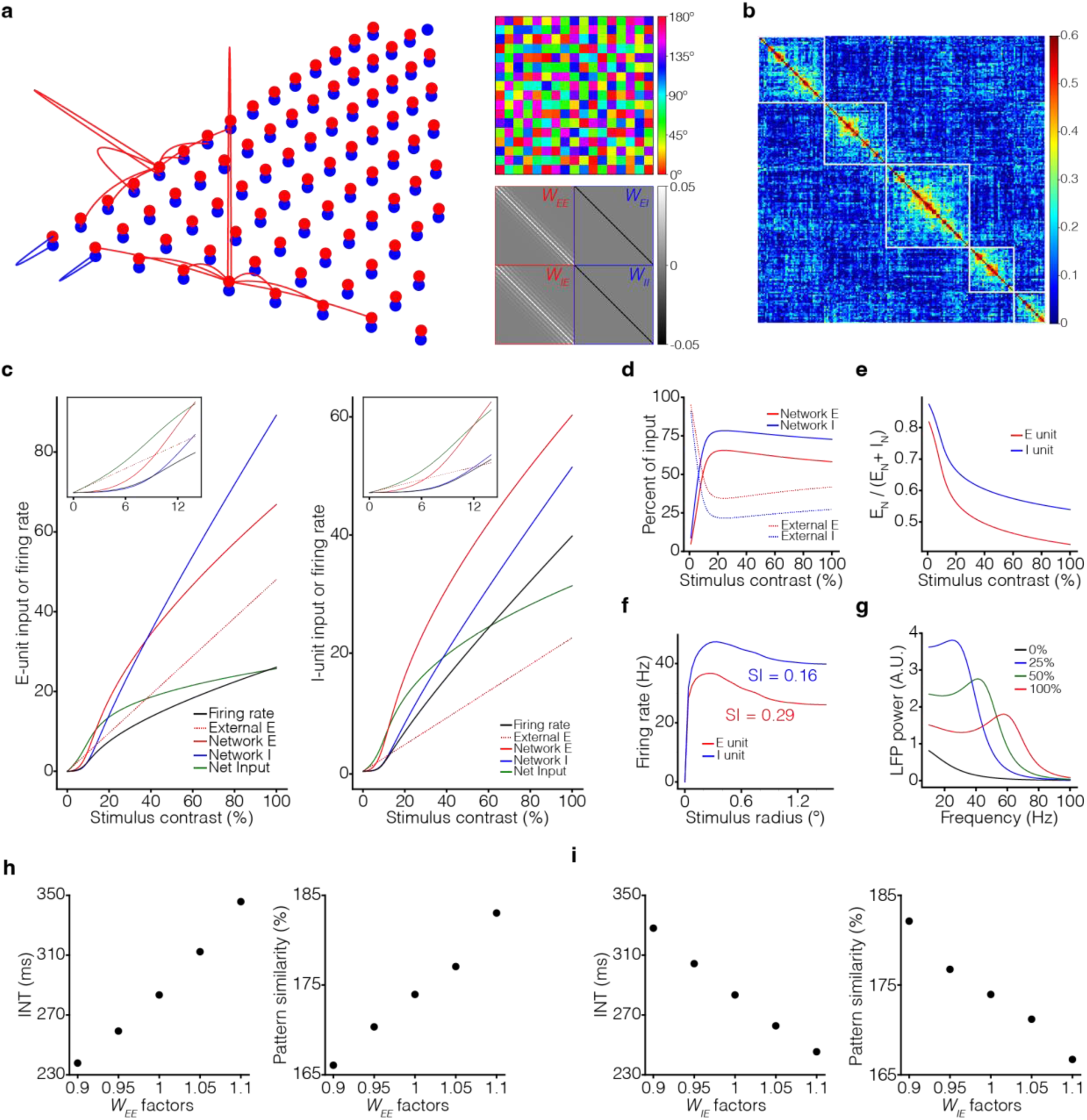
Biophysical model of V1 reproduces pattern similarity and INT deficits through reduced recurrent excitatory connection strengths. **a.** A SSN of V1 was used to investigate pattern similarity alterations previously observed in vivo in three mouse models of schizophrenia-relevant disease mechanisms. The biophysical model (left) consists of 289 excitatory (E; red) and 289 inhibitory (I; blue) neurons equally spaced on a 17-by-17 cm^2^ grid. Connection profiles are displayed for representative neurons. Orientation map (top right) and connectivity matrices (bottom right) of the SSN model. **b.** The similarity matrix from simulated spontaneous activity of the biophysical model demonstrated a highly modular structure (the five clusters determined via k-means clustering are outlined in white), consistent with in vivo observed ensemble activity. **c.** Input to and firing rates of E (left) and I (right) units at stimulus center. With increasing external input stimulus contrast (x axis; dashed lines), network input (E, red and I, blue) transitions from weak to dominating (insets), and substantially cancels external input, so net input (green) grows slowly. Firing rates (black) are proportional to net input squared. **d.** With increasing external input stimulus contrast, input to the network (sum of absolute values of E and I input) is increasingly network-driven (red, input to E unit at stimulus center; blue, input to I unit at stimulus center; dashed, external input; solid, network input). **e.** Network input is increasingly inhibition-dominated with increasing external input stimulus contrast (red, percentage of network input that is excitatory for the E unit at stimulus center; blue, percentage of network input that is excitatory for the I unit at stimulus center). **f.** Size tuning curves of the center E (red) and I (blue) units, at full contrast. E and I firing rates vary non-monotonically with grating size and exhibit surround suppression (suppression indices were 0.29 and 0.16, respectively). **g.** Local field potential (LFP) power-spectra in the center column for external input stimulus contrast *c* = 0%, 25%, 50%, 100% (black, blue, green, and red curves, respectively). The gamma peak frequency increases with increasing external input stimulus contrast. **h.** Simulations over a range of parameter values for connection strengths from E◊E units (*W_EE_*) showing modulation of both INT (left) and pattern similarity (right) as a function of *W_EE_*. Both INT and pattern similarity increase with increasing strength of *W_EE_.* i. Simulations over a range of parameter values for connection strengths from E◊I units (*W_IE_*) showing modulation of both INT (left) and pattern similarity (right) as a function of *W_IE_*. Both INT and pattern similarity decrease with increasing strength of *W_IE_*.

### In vivo test of biophysical model prediction

Finally, given the shared sensitivity of INT and pattern similarity to changes in E→E connection strengths (Figure 3h), when simulating the SSN with varying E→E connection strengths, the biophysical model predicts that INT and pattern similarity should be positively correlated (Figure 4a). This prediction was confirmed in our experimental data where INT and pattern similarity and were correlated both for V1 in mice (t=2.16, p=0.037; Figure 4b) and for whole-brain averages in humans (t=2.25, p=0.025; Figure 4c); results were consistent across mouse models (all interaction effects p>0.281). See Supplement (Supplementary Figure S3) for cortical maps of correlations between INT and pattern similarity across individuals with schizophrenia and non-clinical controls and results for human V1 which showed a similar effect.

**Figure 4.**
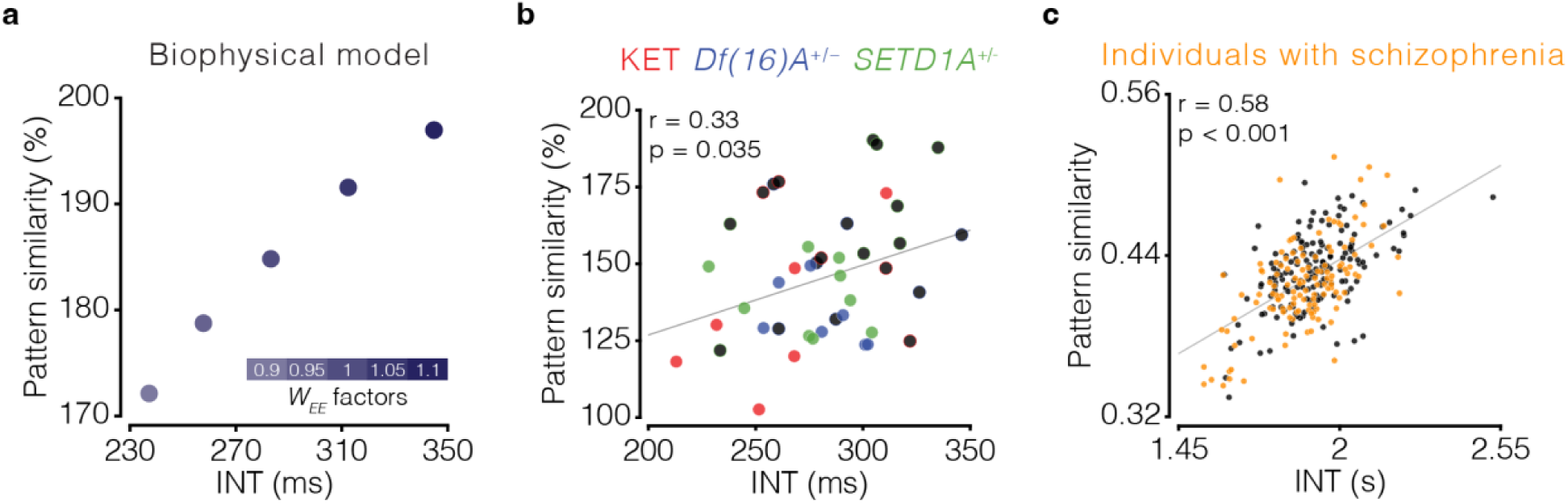
INT and pattern similarity are correlated as predicted by the biophysical model. **a.** When simulating the SSN with varying E→E connection strengths (*W_EE_*), the subsequently calculated INT and pattern similarity are correlated with one another with longer INT and greater pattern similarity resulting from stronger *W_EE_*. This prediction was found to be true in both the mouse 2p-Ca^++^ imaging data (**b**) and the human rs-fMRI data (**c**).

## DISCUSSION

Our findings extend previous work that demonstrated a convergent finding of reduced pattern similarity across three mouse models—two genetic and one pharmacological—related to schizophrenia (35, 36) by identifying convergence across mice and humans, a convergence that applies to two neural phenotypes (pattern similarity and INT). We then used a highly-constrained biophysical model to identify pyramidal neurons as the predominant locus of NMDA-receptor hypofunction and tested a key prediction of this model—correlation between pattern similarity and INT—in both mice and humans.

Although altered excitation-inhibition balance and dysfunctional NMDA receptors are believed to be a core feature of schizophrenia, the exact nature and locus of NMDA-receptor hypofunction is unclear (68–71). We demonstrate that reduced recurrent excitation (i.e. NMDA-receptor hypofunction on pyramidal neurons) accounts for two imaging phenotypes convergent across multiple schizophrenia-relevant mouse models and across mice and humans (Figure 3h)—as well as reduced stimulus-evoked gamma power, which was previously reported across all three mouse models (35, 36) and consistently observed in individuals with schizophrenia (67) (Supplementary Figure S2). This presents possibly the most stringent test to date of a biophysical model applied to schizophrenia and presents a model with greater explanatory power across data types, species, and phenotypes.

Biophysical models are a useful framework for investigating biological mechanisms in psychiatric disorders (72–74) but can suffer from a large parameter search-space where multiple alterations may recapitulate experimentally observed phenotypes. To overcome this challenge, we constrained the parameter search-space by identifying biophysical alterations that recapitulate two observed phenotypes (pattern similarity and INT). Furthermore, we used a SSN model of V1 that has biologically-constrained connectivity, realistic biophysical properties, and recapitulates several neurophysiological aspects of cortical circuits (54–58) including normalization, surround suppression, and contrast dependence of gamma oscillations (Figure 3c–g). With this model, we identified a mechanism with unprecedented explanatory power in that it accounts for mouse and human data within the same model, and it makes predictions that hold in new analyses across species. This is a rare demonstration of an integrative, biologically inspired, approach which opens a path for more effective theory-informed translation.

Our results identify reduced recurrent excitation as an entry point for developing novel therapeutics to treat schizophrenia. Most current pharmacological treatments for schizophrenia are dopamine D2-receptor antagonists that provide limited relief for negative and cognitive symptoms (2–5); although xanomeline–trospium (recently FDA-approved acetylcholine agonist (75)) has reported promising results (76–81). While positive symptoms cause major distress, cognitive symptoms are a hallmark of the disease and are responsible for the greatest reduction in quality of life (82–84). Recurrent excitation facilitates various neuronal phenomena necessary for cognition, including persistent activity during working-memory delay periods (21, 22, 85–87) and evidence accumulation during perceptual decision making (88–91). Thus, future work should investigate relationships between cognition and pattern similarity and INT. Previous work has established a relationship between INT and cognition (92–96), providing compelling evidence for related alterations in schizophrenia, but these remain to be tested and no work has investigated relationships between pattern similarity and INT with cognition in schizophrenia-relevant mouse models. Importantly, future work must test the predictions of the biophysical model by specifically increasing recurrent excitation (e.g., by activation of E→E connections) to see if pattern similarity and INT are subsequently increased. Finally, if a link to cognition is established, targeted increases in recurrent excitation should ameliorate cognitive deficits. Meanwhile, approaches that increase plasticity (97–99) could be effective if a large degree of synapse loss has already occurred (16, 100, 101), and fortifying recurrent excitatory connections during the prodromal stage of schizophrenia could provide a preventative target (101).

However, there are some limitations to consider. Although similar analysis methods were used to estimate INT and pattern similarity across humans and mice, the phenotypes are measured at different scales (voxels containing hundreds of thousands of neurons in humans and single cells in mice). While previous studies have identified convergent properties of INT across units of measurement, similar work should be performed for pattern similarity to further support the translational validity of this neural phenotype. Second, because this was a retrospective secondary data analysis, experimental tests of the theoretically predicted relationship of INT and pattern similarity with cognitive functions dependent upon recurrent excitation could not be performed. In conclusion, although work remains to be done, we believe our convergent translational findings suggest prioritizing the development of treatments for schizophrenia that increase recurrent excitation.

## Data Availability

All data produced in the present study are available upon reasonable request to the authors.

https://schizconnect.org

https://openfmri.org/dataset/ds000030/

## Acknowledgements

This work was supported by the National Institute of Mental Health under award R01MH134973 to Dr. Kenneth Wengler. BrainGluSchi: data were downloaded from the COllaborative Informatics and Neuroimaging Suite Data Exchange tool (COINS;http://coins.mrn.org/dx) and data collection was funded by NIMH R01MH084898-01A1. COBRE: Data was downloaded from the COllaborative Informatics and Neuroimaging Suite Data Exchange tool (COINS; http://coins.mrn.org/dx), data collection was performed at the Mind Research Network, and funded by a Center of Biomedical Research Excellence (COBRE) grant 5P20RR021938/P20GM103472 from the NIH to Dr. Vince Calhoun. NMorphCH: data were obtained from the Neuromorphometry by Computer Algorithm Chicago (NMorphCH) dataset (http://nunda.northwestern.edu/nunda/data/projects/NMorphCH); the investigators within NMorphCH contributed to the design and implementation of NMorphCH and/or provided data but did not participate in analysis or writing of this report; data collection and sharing for this project was funded by NIMH grant R01MH056584. UCLA: data was obtained from the OpenfMRI database (its accession number is ds000030) and data collection was funded by the Consortium for Neuropsychiatric Phenomics (NIH Roadmap for Medical Research grants UL1-DE019580, RL1MH083268, RL1MH083269, RL1DA024853, RL1MH083270, RL1LM009833, PL1MH083271, and PL1NS062410).

