## Supplementary Material for "Convergent multidimensional neural phenotypes in human patients and schizophrenia-relevant mouse models identify reduced recurrent excitation in cortical circuits"

### SUPPLEMENTARY MATERIALS

### Methods

#### *Schizophrenia Combined Dataset*

All human data presented here come from datasets published in Wengler et al. (2020)[30]. As previously described[30], T1w images and rs-fMRI data were obtained for 331 non-clinical control subjects and 254 individuals diagnosed with either schizophrenia (N = 241) or schizoaffective disorder (N = 13) from four publicly available datasets. Three of these datasets were from the SchizConnect repository (BrainGluSchi[37], COBRE[38,39], and NmorphCH[40]) and one was from the OpenfMRI repository (UCLA[41]). Data that survived a quality-control check (~95%) and motion-censoring check (~64%) included 140 individuals with schizophrenia and 225 controls. The quality control check consisted of visual inspection of the spatially normalized images. The motion censoring check consisted of determining if there were sufficient degrees of freedom after motion censoring to perform nuisance variable regression. A subset of 158 controls was then selected that matched individuals with schizophrenia on gender and age. To minimize scanner- and site-related differences we excluded subjects if the signal-to-noise ratio (SNR) was less than 100 for any of the standard regions-of-interest[102]. The final sample after quality-control checks consisted of 127 individuals with schizophrenia and 152 age- and gender-matched controls (**Table S1**).

The rs-fMRI data were collected for each subject during an eyes-open-on-fixation session with the following scanning parameters: TR = 2000 ms (except for NmorphCH, where TR = 2200 ms); time points (BrainGluSchi/COBRE/NmorphCH/UCLA) = 160/145/318/147; spatial resolution (mm) = 3.5×3.5×3.5/3.5×3.5×3.5/4×4×4/3×3×3. Data were preprocessed using the AFNI afni_proc.py function[103]. The following steps were performed: (1) removal of the first five volumes with the 3dTcat function; (2) slice-timing correction; (3) motion correction; (4) 12-parameter affine registration of the rs-fMRI images to the T1w image; (5) spatial normalization of rs-fMRI images to MNI152_ICBM2009a_nlin volume space using nonlinear warping via the T1w image; (6) single-interpolation resampling of rs-fMRI images combining motion correction and spatial normalization. The preprocessed rs-fMRI data were the postprocessed with the following steps: (1) regression of white-matter signal, cerebrospinal-fluid signal, global-brain signal, and the 6 motion parameters along with their first derivatives; (2) bandpass filtering in the 0.01–0.1 Hz range; (3) motion censoring to remove volumes with framewise displacement (FD) [104] greater than 0.3 mm along with the volumes directly preceding and following that volume; (4) spatial smoothing with a 4 mm full-width-at-half-maximum Gaussian kernel; (5) Lomb-Scargle interpolation of censored frames.

INT maps were calculated from the postprocessed rs-fMRI data as previously described[30]. Briefly, the autocorrelation function of the rs-fMRI signal at each voxel was estimated and the sum of the autocorrelation coefficients during the initial positive period was calculated. This initial positive period included all timepoints from the first timepoint (lag one) until the timepoint immediately preceding the first lagged timepoint with a non-positive autocorrelation coefficient. To adjust for differences in temporal resolution, the sum was multiplied by the repetition time (TR) of the rs-fMRI data. This product was used as an index for INT. Voxelwise INT maps were parcellated into 180 cortical parcels using HCP-MMP1.0 in volumetric space averaging across the left and right hemispheres.

Pattern similarity maps were calculated from the postprocessed rs-fMRI data according to methods similar to those used for 2p-$Ca^{2+}$ imaging data with neural events being defined exactly as in co-activation pattern analyses of rs-fMRI data. The following steps were performed separately for each of the 180 cortical parcels to estimate pattern similarity values for each parcel: (1) rs-fMRI signal from each voxel within the parcel was temporally z-scored; (2) neural events were defined as frames having a z-score greater than 1, with frames not defined as neural events were set to zero (i.e. event thresholding); (3) high coactivity frames were determine by selecting the 30 non-neighboring frames with the highest average z-scored and thresholded signal across voxels; (4) t-distributed stochastic neighbor embedding (t-SNE) analysis to reduce the high dimensional high coactivity frames (1 dimension per voxel) to a low dimensional subspace (3-dimesions) where activity patterns are grouped by their nearest neighbors; (5) k-means clustering analysis on this reduced dataset with k=2; (6) reapplied the clustering solution (i.e. state-cluster memberships) back to the original high coactivity frames activations (pre-t-SNE), and calculated average within-cluster cosine similarity values ($c_{WC}$). The $c_{WC}$ value within the dominant cluster (i.e. the cluster with higher $c_{WC}$) was taken as the pattern similarity.

The parcellated INT and pattern similarity maps were then harmonized using ComBat[45-49] controlling for age and sex. The harmonization parameters were estimated using data from the non-clinical controls and subsequently applied to all subjects to minimize the potential removal of biological variability between non-clinical controls and individuals with schizophrenia[50-52].

***Mouse Models of Schizophrenia-Relevant Disease Mechanisms Combined Dataset***

All animal work was carried out under IACUC approved protocols at Columbia University. All mouse data presented here come from image-processed datasets published in Hamm et al. (2017)[35] and Hamm et al. (2020)[36]. All analyses in focused on brain activity collected in a darked room in awake mice in the absence of structured or changing visual input, which we refer to herein as "spontaneous activity." Pattern similarity analyses match those described in Hamm et al. (2020)[36], and data from Hamm et al. (2017)[35] was re-analysed to match those procedures. INT was not previously analyzed in either of these datasets. The surgical, experimental, and data processing steps are identical to those works[35,36] and are only detailed briefly here.

Three previously published mouse datasets were used: a group of mice given 100mg/kg/day ketamine (n=6; 4 males) or saline (n=5; 3 males) continuously for a week via a subcutaneous minipump (from Hamm et al., 2017); a group of *Df(16)A*$^{+/-}$ mice (mirroring the 22q11.2 microdeletion syndrome; n=7, all male) or their littermate/cagemate wild-type (WT) controls (n=7, all male; from Hamm et al., 2017[35]); or a group of *SETD1A*$^{+/-}$ mice (n=9; all male) or their littermate/cagemate WT controls (n=8; all male; from Hamm et al., 2020[36]). One saline mouse and one WT control from the *SETD1A*$^{+/-}$ cohort were excluded from the originally published datasets due to insufficient signal to noise ratio for calculating INT using previously established criteria[95,96]. All mice were aged between P84 and P180 during recordings.

In order to image the activity of neurons, layer 2/3 (250 um deep) in putative monocular primary visual cortex was injected with 700 nl of AAV1/Syn:GcaMP6s (University of Pennsylvania Vector Core; diluted 1:1 with PBS buffer) over a 5 min period. Approximately two weeks after virus injection, a titanium head plate was attached to the as skull with dental cement. On the day of imaging, a small circle (approximately 2 mm in diameter) was thinned or removed over the left V1 centered just anterior to the injection site. The mouse was then allowed to wake up and was transferred to the wheel.

Two-photon calcium imaging was carried out in a dark room in the absence of visual stimulation. Mice were awake and head-fixed under the microscope on a wheel. Locomotion was measured with an infrared LED-photodarlington pair attached to the treadmill and did not differ between any groups (see Hamm et al., 2017, 2020[35,36]). Bouts during locomotion were typically only present in ≈5% of frames and were excluded from further analyses. The activity of cortical neurons was recorded by imaging fluorescence (F) changes under a two-photon microscope (Bruker, Billerica, MA) excited with a Ti:Sapphire laser (Chameleon Ultra II [Coherent, Santa Clara, CA] or Mai Tai HP Deep See [Spectra Physics, Santa Clara, CA]) tuned at 940 nm and scanned with resonant galvometers through a ×20 (numerical aperture 0.9) water immersion objective (Olympus, Center Valley, PA). Resonant-galvo (*SETD1A*$^{+/-}$ data) or galvo-galvo (ketamine and *Df(16)A*$^{+/-}$ data) scanning and image acquisition were controlled by PrairieView (Bruker, Billerica, MA) software. For the *SETD1A*$^{+/-}$ recordings, images were collected at ≈10 frames per second for 256×256 pixels, 200–225 μm beneath the pial surface. For the ketamine and *Df(16)A*$^{+/-}$ recordings, images were collected at ≈3.4 frames per second. Imaging consisted of a visual stimulation condition (15 minutes), followed by 20–40 minutes of awake rest in a dark room with the monitor off, followed by a second visual stimulation.

The raw images were processed to correct translational brain motion artifacts using the MOCO plugin for ImageJ. Then, cell regions of interest (ROIs) were detected semi-automatically (locally constrained PCA) and their corresponding fluorescence traces were visually inspected. Cells with faint, sparse, or largely atypical calcium transients were excluded from further analysis. We carried out a “halo” subtraction, eliminating contamination of neighboring pixels and ambient room light. The remaining traces were then filtered with a 1-second lowess envelope and the first discrete derivative was scored as Δf (for the INT analysis) or within-cell maximum normalized Δf for population analyses (for the population pattern similarity analysis).

Individual neuron timescales were calculated by fitting the Δf timeseries to an exponential decay function and the mouse’s INT was taken as the median of the decay rates across the imaged neurons.

To analyze the pattern similarity of spontaneous ensemble activations during resting state, we carried out a set of steps on each dataset. The approach is identical to what is reported in Hamm et al. (2020)[36] and was reapplied to the dataset in Hamm et al. (2017)[35]. First, a bootstrap approach was carried out on individual datasets in order to determine what characterized a true “ensemble activation” peak. We identified high coactivity frames that represented the peak of the preceding and subsequent 1.5 seconds and exceeded the 99.9$^{th}$ percentile of coactivity from time-shuffled datasets. Each dataset exhibited between 150 and 250 peak “ensembles activations” during the first 20 minutes ongoing activity.

Next, we sought to determine which “ensemble activations” showed population level patterns of activation which repeated over the rest period, suggesting attractor states in the ongoing activity. We first carried out a t-distributed stochastic neighbor embedding (t-SNE) analysis[105] to reduce the high dimensional ensemble activation datasets where activity patterns are grouped by their nearest neighbors. This resulted in a data set of states×dimensions for each mouse.

In the next step, we carried out a k-means clustering analysis on this reduced dataset (state×dimension). Our past work shows that our results were robust across multiple values of k (i.e. across multiple numbers of clusters). For simplicity, we fixed the number of clusters (i.e. the value of k) to the number of neurons in the dataset divided by 33 (i.e. 3 clusters per 100 neurons) as previously described (see e.g. Figure 2J from Hamm et al., 2020). We reapplied these solutions (i.e. state-cluster memberships) back to the original ensemble activations (pre-t-SNE; state×cells), and calculated average within-cluster similarity values using Pearson correlations. To normalize these values, each r-value was divided by the average unclustered correlation value across all ensemble activations. This normalized number—our pattern similarity—represents the degree to which ensemble activations within the same "cluster" were similar to one another and unique from other activation states.

***Biophysical Model***

We utilized the stabilized supralinear network (SSN)[54-58] with different synaptic receptor types[56]. The SSN is highly constrained by biological properties and has been shown to recapitulate many features of the V1. Here, we implemented a retinotopically-structured SSN model of V1 that models the cortex as a two-dimensional grid with an *E* and *I* sub-population (corresponding to SSN units) at each grid location, corresponding to a cortical column. The grid two-dimensional grid covered an area of 6.4 mm × 6.4 mm and contained 17 × 17 evenly spaced cortical columns (each containing and *E* and *I* sub-population) such that the total number of *E* and *I* populations is 289 (N=578). We account for the distinct dynamics of currents through different synaptic receptor channels: AMPA, $GABA_A$ (henceforth GABA), and NMDA. The dynamical state of the network, in a network of N neurons, is given by the N-dimensional vector of inputs $\boldsymbol{h}_t$, which evolves according to the dynamical system:

$$\tau^{\alpha}\frac{d}{dt}\boldsymbol{h}_t^{\alpha} = -\boldsymbol{h}_t^{\alpha} + W^{\alpha}\boldsymbol{r}_t + \boldsymbol{I}_t^{\alpha} \qquad (1)$$

where $\boldsymbol{r}_t$ is the vector of firing rates, $W^{\alpha}\boldsymbol{r}_t$ and $\boldsymbol{I}_t^{\alpha}$ denote the recurrent and external inputs to the network mediated by receptor $\alpha$ ($\alpha \in \{\mathrm{A} = \mathrm{AMPA}, \mathrm{G} = \mathrm{GABA}, \mathrm{N} = \mathrm{NMDA}\}$), respectively, and $W^{\alpha}$ are N × N matrices denoting the contributions of different receptor-types to recurrent connectivity weights; the total recurrent connectivity weight matrix is thus given by $W \equiv \sum_{\alpha} W^{\alpha}$. To close the system for the dynamical variables $\boldsymbol{h}_t^{\alpha}$, we have to relate the output rate of a neuron to its total input current. Here we use the instantaneous approximation to the input-output (I/O) transfer function of neurons[106,107] such that:

$$\boldsymbol{r}_t = F\left(\boldsymbol{h}_t^{total}\right) = k\left[\sum\nolimits_{\alpha} \boldsymbol{h}_t^{\alpha}\right]_+^n \quad (2)$$

where the I/O function $F(\cdot)$ acts element-wise on its vector argument. As in the original SSN[58], we take this I/O transfer function to be a supralinear rectified power-law, which is the essential ingredient of the SSN: $F(v) \equiv k[v]_+^n$, where k is a positive constant, n > 1 (corresponding to supralinearity), and $[x]_+ \equiv \max(0, x)$ denotes rectification. While the I/O function of biological neurons saturates at high firing rates (e.g., due to refractoriness), throughout the natural dynamic range of cortical neurons firing rates stay relatively low. In fact, in V1 neurons the relationship between the firing rate and the mean membrane potential (an approximate surrogate for the neuron's net input) shows no saturation throughout the entire range of firing rates driven by visual stimuli, and is well approximated by a supralinear rectified power-law[108,109]. Since AMPA and NMDA only contribute to excitatory synapses, and GABA only to inhibitory ones, in general the $W^{\alpha}$ have the following block structure:

$$W^{\mathrm{A}} = \begin{pmatrix} W_{EE}^{\mathrm{A}} & 0 \\ W_{IE}^{\mathrm{A}} & 0 \end{pmatrix}, \quad W^{\mathrm{N}} = \begin{pmatrix} W_{EE}^{\mathrm{N}} & 0 \\ W_{IE}^{\mathrm{N}} & 0 \end{pmatrix}, \quad W^{\mathrm{G}} = \begin{pmatrix} 0 & -W_{EI}^{\mathrm{G}} \\ 0 & -W_{II}^{\mathrm{G}} \end{pmatrix} \quad (3)$$

For simplicity, we further assumed that the fraction of NMDA and AMPA is the same in all excitatory synapses. In this case all $W^{\alpha}$ can be written in terms of the four blocks of the full connectivity matrix $W \equiv \sum_{\alpha} W^{\alpha}$:

$$W^{\mathrm{N}} = \frac{\rho_{\mathrm{N}}}{1-\rho_{\mathrm{N}}} W^{\mathrm{A}} = \rho_{\mathrm{N}} \begin{pmatrix} W_{EE} & 0 \\ W_{IE} & 0 \end{pmatrix}, \quad W^{\mathrm{G}} = \begin{pmatrix} 0 & -W_{EI} \\ 0 & -W_{II} \end{pmatrix} \tag{4}$$

where the scalar $\rho_{\mathrm{N}}$ is the fractional contribution of NMDA to excitatory synaptic weights. The external input ($\boldsymbol{I}_t^{\alpha}$) has the following form:

$$\boldsymbol{I}_t^{\alpha} = \boldsymbol{I}_{Ext}^{\alpha} + \eta_t^{\alpha} \tag{5}$$

where $\boldsymbol{I}_{Ext}^{\alpha}$ represents the feedforward background or stimulus drive to the network (by a steady time-independent stimulus) and scales with the contrast of the visual stimulus, and $\eta_t^{\alpha}$ represents the stochastic noise input to the network. For the resting-state condition (i.e. spontaneous activity), $\boldsymbol{I}_{Ext}^{\alpha}$ was determined such that the excitatory and inhibitory cells have firing rates of 1.5 Hz and 3 Hz, respectively[57]. Given that external inputs to cortex are excitatory, we assumed that $\eta_t^{\alpha}$ was zero for α = GABA. We took $\eta_t^{\alpha}$ to have spatially correlated and identically distributed components across our sub-population units, and took it to be temporally correlated pink noise with zero mean and unit variance. The degree of noise correlation between neurons was decayed as an exponential function of distance between neurons with a decay constant of 0.4 mm (one grid interval)[59-61]. For the visual-stimulus condition, we used full-field gratings with varying strengths of contrast (see Holt et al., 2024[56] for details).

The 2×2 (full) connectivity matrix is parametrized by the four parameters $J_{ab}$ $(a, b \in \{E, I\})$ as follows:

$$W = \begin{pmatrix} J_{EE} & -J_{EI} \\ J_{IE} & -J_{II} \end{pmatrix} \tag{6}$$

We indexed the neurons by their E/I type and retinotopic location. We parametrized the recurrent connection weight from the pre-synaptic E and I units at location **y** with preferred orientation $\theta_{\mathrm{y}}$ to the type $a$ $(a \in \{E, I\})$ post-synaptic unit at location **x** with preferred orientation $\theta_{\mathrm{x}}$ by:

$$W_{\mathrm{x},a|\mathrm{y},E} \propto J_{a,E}\left[\lambda_{a,E}\delta_{\mathrm{x,y}} + (1-\lambda_{a,E})e^{-\frac{\|\mathrm{x}-\mathrm{y}\|}{\sigma_{a,E}} - \frac{(\theta_{\mathrm{x}}-\theta_{\mathrm{y}})^2}{2\sigma_{\theta,}^2}}\right] \quad (6)$$

for excitatory projections, and

$$W_{\mathrm{x},a|\mathrm{y},I} \propto J_{a,I}e^{-\frac{(\mathrm{x}-\mathrm{y})^2}{2\sigma_{a,I}^2} - \frac{(\theta_{\mathrm{x}}-\theta_{\mathrm{y}})^2}{2\sigma_{\theta,}^2}} \quad (7)$$

for inhibitory projections[110-112]. We are using proportionality instead of equal signs in the Eqs. 6 and 7, because a normalization was done such that the total weight of each type received by a unit was given by the corresponding $J_{a,b}$ (independent of the $\sigma_{a,b}$ and $\lambda_{a,E}$ parameters). Recurrent connectivity was thus parametrized by the 2x2 matrices $J_{a,b}$ and $\sigma_{a,b}$, $\sigma_{\theta},$ the two $\lambda_{a,E}$, and the NMDA fraction, $\rho_{\mathrm{N}}$, 12 parameters in total. For $\sigma_{I,I}$ and $\sigma_{E,I}$ we used values small compared to the distance between neighboring columns (0.4 mm) so that inhibition was effectively local (i.e. intra-columnar). For $\sigma_{\theta}$, we used a value estimated from mouse V1 data[113].

Biophysical model simulations were run using the forward Euler method with a time step of 1 ms and a total simulation time of 10 minutes. Simulations were performed while independently varying one of each of the connectivity strengths (*$W_{EE}$, $W_{IE}$, $W_{EI}$,* or *$W_{II}$*) while keep all other parameters fixed. The connectivity strengths were varied by factors ranging from 0.9 to 1.1 (i.e. 10% decrease to 10% increase) in steps of 5% (i.e. 5 separate values), for a total of 20 separate model simulations (i.e. 5 connectivity values for each of the 4 connectivity parameters).

**Table S2** lists the values of parameters and parameter ranges used in simulations. Parameter values were chosen to match those in Holt et al., 2024[56], and we provide a brief description of choices here. The values for $J_{ab}$, $g_a$, $\sigma_{aE}$, $\lambda_{aE}$, and $\rho_N$ were chosen by random sampling such that the network would exhibit the local contrast-dependence of the gamma peak together with strong surround suppression. Note that $g_a$ represents the feedforward current per

1% contrast to excitatory or inhibitory cells. This parameter is not relevant in the current simulations because we only simulated the network in the resting-state condition (i.e. no external stimulus), but was used to derive other network parameters as outlined below. Parameters were sampled from wide biologically plausible ranges. To determine the ranges for the recurrent and feedforward weights, rough biological estimates for the recurrent E and I weights (i.e. $J_{aE}$ and $J_{aI}$, respectively, for $a \in \{E, I\}$), as well as the (excitatory) feedforward weights ($g_E$ and $g_I$) were made. Parameters were then independently varied controlling each type of weight between 0.5 to 1.5 times those estimates. The effect of feedforward inputs on membrane voltage was estimated using measurements in cats and mice[114-118] (see Ahmadian and Miller, 2021[119] for a review and discussion of these measurements). Based on these measurements, the maximum feedforward input, achieved for 100% contrast, was estimated to be on the order of the rest to threshold distance, which is around 20 mV[120]. This yields $g_a = 20\ \mathrm{mV}/s$ per 1% contrast. The $J_{ab}$ parameters measure the total synaptic weight, which biologically is given by a unitary excitatory or inhibitory (depending on *b*) post-synaptic potential (EPSP or IPSP) times the total number of pre-synaptic V1 neurons, $K_b$, of type *b*. Based on anatomical measurements for sensory cortex (reviewed in Ahmadian and Miller, 2021[119]), the effective $K_E$ was estimated to be ~400. Based on electrophysiological measurements, the median EPSP amplitude was assumed to be ~0.5 mV. This yields $J_{aE} = 0.5 \times 400 = 200\ \mathrm{mV}$. For unitary IPSP amplitude, the same value of 0.5 mV was used, but assumed half as many inhibitory pre-synaptic inputs, due to the smaller number of inhibitory cells in the circuit, thus yielding $J_{aI} = J_{aE}/2$. The parameters $\lambda_{EE}$ and $\lambda_{IE}$ quantifying the intra-columnar excess connectivity were sampled from the interval [0.25, 0.75]. The parameters $\sigma_{EE}$ and $\sigma_{IE}$ quantifying the length-scale (range) of the long-range components of excitatory recurrent connections were sampled between 150 μm and 600 μm. Finally, the recurrent V1 excitatory synapses were assumed to be dominated by AMPA, rather than NMDA, and $\rho_N$ was therefore sampled in the interval [0.3, 0.5]. All parameters were sampled uniformly and independently over their ranges, except for enforcement (by sample rejection) of three inequality constraints: $J_{EI}J_{IE} > J_{EE}J_{IE}$, $J_{II}g_E > J_{EI}g_I$, and $\sigma_{IE} > \sigma_{EE}$. Previous work has shown that the first inequality promotes stability (almost a necessary condition)[54,121], the second inequality ensures that the network is not too strongly inhibition-dominated such that excitatory rates become too small[54,121]. The last inequality is necessary for obtaining considerable surround suppression[58].

The simulated firing rates from each of the 289 excitatory neurons were used to calculate INT and pattern similarity (to match the 2P-$Ca^{2+}$ data that imaged layer 2/3 pyramidal neurons). The first minute of simulated firing rates were discarded and the simulated firing rates were downsampled to a temporal resolution of 10 Hz. The model's INT was estimated using the same procedure as the calcium imaging data. The model's pattern similarity was estimated using the same procedure as the calcium imaging data using the top 200 timepoints (determined via bootstrapping of the unperturbed model) and k=5 (determined via maximizing the silhouette index of the ensemble data from the unperturbed model).

### Supplemental Figures

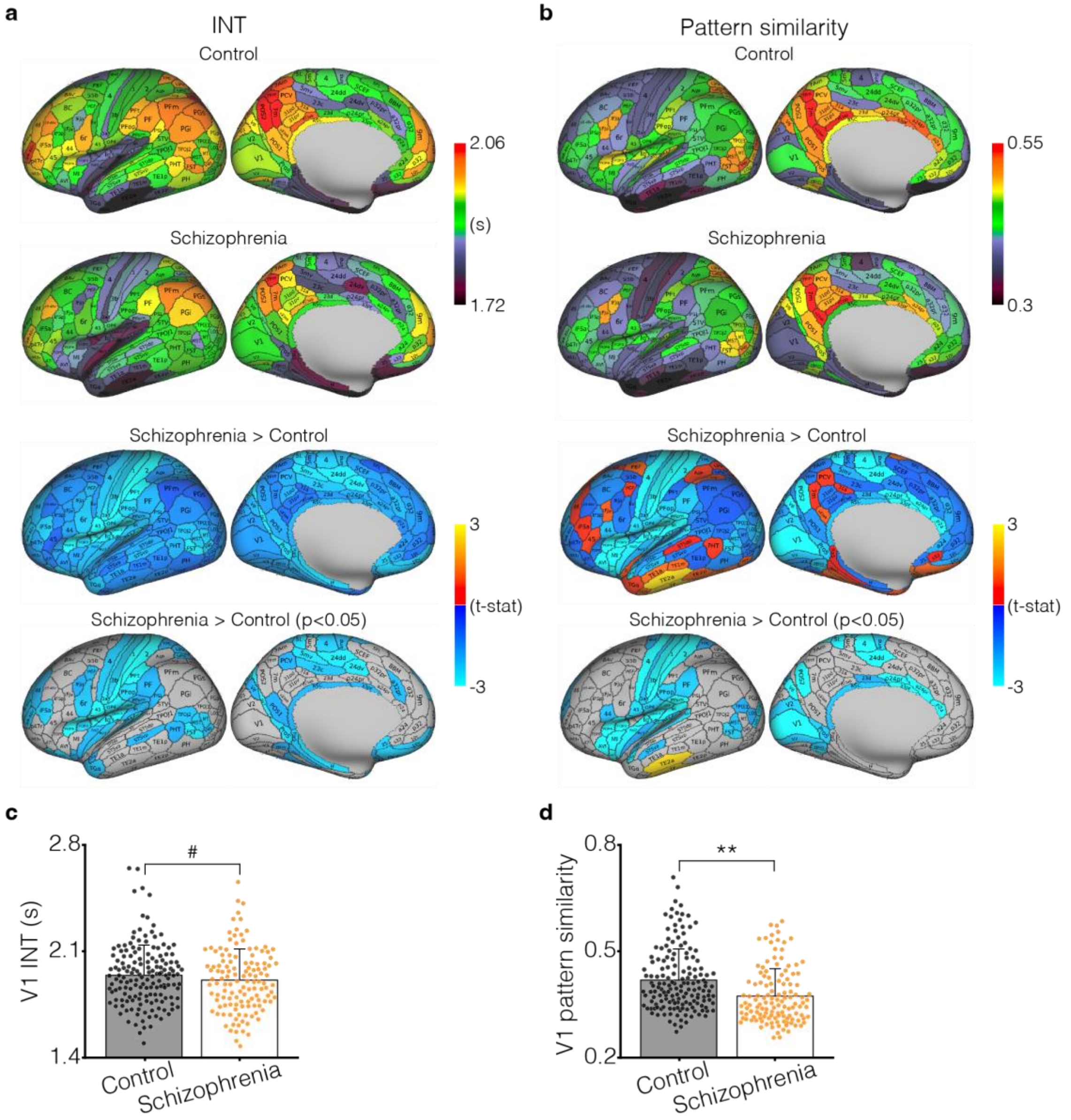


**Figure S1. Whole-brain maps of INT and pattern similarity with related statistical tests, and V1-specific effects. a.** Whole-brain maps of INT averaged across non-clinical controls (top) and individuals with schizophrenia (middle-top) with the t-statistic map without thresholding (middle-bottom) and with thresholding for significant effects (bottom) for the contrast of individuals with schizophrenia greater than non-clinical controls (i.e. blue denotes regions where INT was shorter in the schizophrenia group compared to the controls). **b.** Whole-brain maps of pattern similarity averaged across non-clinical controls (top) and individuals with schizophrenia (middle-top) with the t-statistic map without thresholding (middle-bottom) and with thresholding for significant effects (bottom) for the contrast of individuals with schizophrenia greater than non-clinical controls (i.e. blue denotes regions where pattern similarity was shorter in the schizophrenia group compared to the controls) **c.** INT is trend-level shorter in V1 of individuals with schizophrenia compared to non-clinical controls. **d.** Pattern similarity is significantly lower in V1 of individuals with schizophrenia compared to non-clinical controls. #, $p<0.1$; **$p<0.01$.

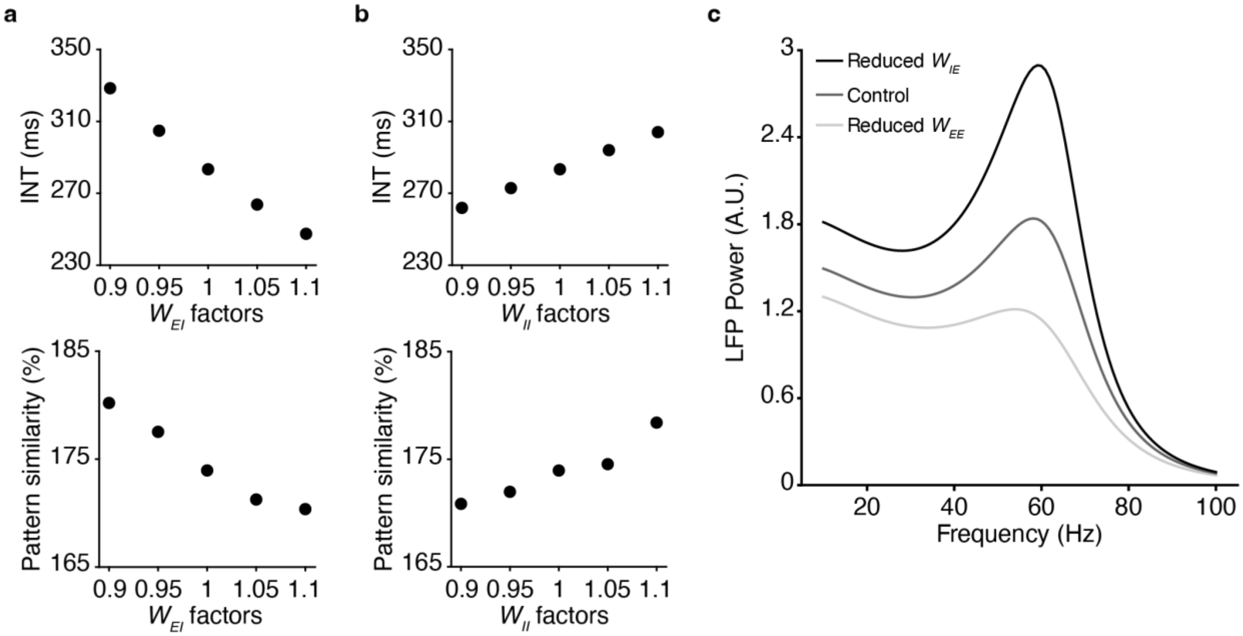


**Figure S2. Biophysical model simulations for inhibitory connection strengths and recapitulation of reduced stimulus-evoked gamma power. a.** SSN model simulations over a range of parameter values for connection strengths from I→E units ($W_{EI}$) showing modulation of both INT (top) and pattern similarity (bottom) as a function of $W_{EI}$. Both INT and pattern similarity decrease with increasing strength of $W_{EI}$. **b.** Simulations over a range of parameter values for connection strengths from I→I units ($W_{II}$) showing modulation of both INT (top) and pattern similarity (bottom) as a function of $W_{II}$. Both INT and pattern similarity increase with increasing strength of $W_{II}$. **c.** Reduced stimulus-evoked gamma power is a consistently observed phenotype in individuals with schizophrenia. Simulations with reduced connection strengths from E→E units ($W_{EE}$; 5% reduction) recapitulate reduced stimulus-evoked gamma power while simulations with reduced connection strengths from E→I units ($W_{IE}$; 5% reduction) produce increased stimulus-evoked gamma power, further supporting pyramidal neurons as the predominant locus of NMDA-receptor hypofunction in schizophrenia.

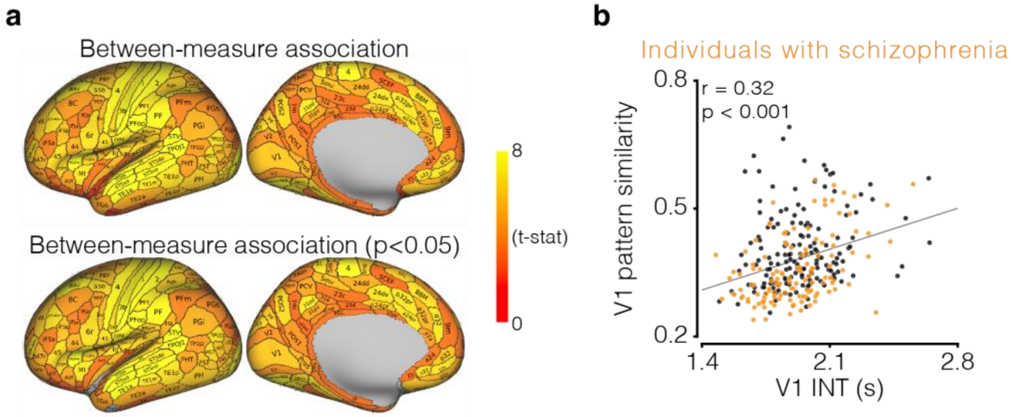


**Figure S3. Whole-brain maps of the relationship between INT and pattern similarity, and V1-specific effect. a.** Whole-brain t-statistic maps without thresholding (top) and with thresholding for significant effects (bottom) for the relationship between INT and pattern similarity (note that only three brain regions are not significant). **b.** Scatter plot showing the relationship between INT and pattern similarity within V1.

### Supplemental Tables

#### Table S1. Human participant characteristics

| Variable | Non-clinical controls | | | | | Individuals with schizophrenia | | | | |
|---|---|---|---|---|---|---|---|---|---|---|
| | BGS | COBRE | NMCH | UCLA | All | BGS | COBRE | NMCH | UCLA | All |
| N | 24 | 42 | 25 | 67 | 158 | 40 | 31 | 26 | 30 | 127 |
| Age, mean (SD), y | 36.0 (13.1) | 33.9 (10.4) | 29.8 (7.2) | 32.3 (8.5) | 32.9 (9.7) | 31.1 (12.6) | 30.5 (11.9) | 30.5 (6.2) | 35.0 (8.9) | 31.8 (10.6) |
| Male sex, No. (%) | 22 (92) | 34 (81) | 16 (64) | 54 (81) | 126 (80) | 38 (95) | 26 (84) | 19 (73) | 22 (73) | 105 (83) |
| Framewise Displacement*, mean (SD), mm | 0.15 (0.04) | 0.14 (0.05) | 0.14 (0.11) | 0.10 (0.04) | 0.13 (0.06) | 0.16 (0.06) | 0.17 (0.06) | 0.12 (0.06) | 0.13 (0.04) | 0.15 (0.06) |
| Percent of censored frames, mean (SD) | 8.91% (6.73%) | 9.94% (8.12%) | 9.59% (9.81%) | 4.45% (5.79%) | 7.40% (7.69%) | 9.21% (7.21%) | 11.09% (7.89%) | 9.18% (7.35%) | 9.75% (8.25%) | 9.79% (7.61%) |

*Framewise Displacement values were estimated after motion-scrubbing
BGS, BrainGluSchi; NMCH, NMorphCH; SD, standard deviation

#### Table S2. Parameters used for SSN biophysical model simulations.

| Parameter | Value | Unit | Description |
|---|---|---|---|
| $n$ | 2 | - | power-law I/O exponent |
| $k$ | 0.0194 | $mV^{-2} \cdot ms$ | power-law I/O pre-factor |
| $\tau_{corr}$ | 5 | ms | noise correlation time |
| $\tau_{AMPA}$ | 5 | ms | AMPA decay time |
| $\tau_{GABA}$ | 7 | ms | $GABA_A$ decay time |
| $\tau_{NMDA}$ | 100 | ms | NMDA decay time |
| $\rho_N$ | 0.39 | - | NMDA share of excitation |
| $J_{EE}$ | 124 ± 10% | mV | total E → E connection weight |
| $J_{IE}$ | 116 ± 10% | mV | total E → I connection weight |
| $J_{EI}$ | 103 ± 10% | mV | total I → E connection weight |
| $J_{II}$ | 59.3 ± 10% | mV | total I → I connection weight |
| $\lambda_{EE}$ | 0.72 | - | locality of E → E connections |
| $\lambda_{IE}$ | 0.70 | - | locality of E → I connections |
| $\sigma_{EE}$ | 0.296 | mm | range of E → E connections |
| $\sigma_{IE}$ | 0.554 | mm | range of E → I connections |
| $\sigma_{EI}$ | 0.09 | mm | range of I → E connections |
| $\sigma_{II}$ | 0.09 | mm | range of I → I connections |
| $\sigma_\theta$ | 45 | ° | orientation tuning width of connections |
| $N_{col}$ | $17^2$ | - | number of cortical columns |
| $L$ | 6.4 | mm | retinotopic network width |
| $\Delta x$ | 0.4 | mm | cortical column width |
